# Prediction model of hypertensive disorders of pregnancy based on home blood pressure monitoring

**DOI:** 10.1101/2025.05.28.25328541

**Authors:** Makito Oku, Shuntaro Ohno, Hirohito Metoki, Eri Arai, Mika Ito, Arihiro Shiozaki, Akitoshi Nakashima, Noriyuki Iwama, Rie Tsuchida, Satoshi Yoneda, Noriko Yoneda, Sayaka Tsuda, Michihiro Satoh, Takahisa Murakami, Shingo Tanigawara, Kazuhiko Hoshi, Kohei Tanaka, Masaaki Yamada, Shigeru Saito

## Abstract

**Background:** Hypertensive disorders of pregnancy (HDP) cause adverse maternal and fetal outcomes, and establishing an early prediction method for HDP is needed. Existing methods based on serum markers require blood sampling. Therefore, we aimed to develop and validate a noninvasive HDP prediction model based on home blood pressure monitoring (HBPM).

**Methods:** In a development cohort, HBPM data from 443 pregnant women including 65 HDPs were divided into training data (n=365) and test data (n=78) to develop a logistic regression-based prediction model. Normal blood pressure variations depending on season and gestational age were subtracted. At each time point, four features were calculated for the last four weeks of data: average systolic blood pressure (SBP), average diastolic blood pressure (DBP), correlation coefficient between SBP and DBP, and upward trend of SBP against day. In a validation cohort, HBPM data from 264 pregnant women including 33 HDPs were collected prospectively and used to validate the model.

**Results:** The area under the receiver operating characteristic curve was 0.949, 0.884, and 0.845 for the training, test, and validation data, respectively. Sensitivity, specificity, positive predictive value, and negative predictive value were 0.923, 0.888, 0.387, and 0.993 for the training data; 0.667, 0.897, 0.867, and 0.729 for the test data; 0.758, 0.766, 0.316, and 0.957 for the validation data.

**Conclusions:** Our HDP prediction model based on HBPM showed high negative predictive values and may contribute to reducing medical consultations.

**Clinical Perspective:** *What Is New?:*

- A prediction model for hypertensive disorders of pregnancy based on home blood pressure monitoring was developed, and its performance was validated in a separate cohort.

*What Are The Clinical Implications?:*

- The proposed method does not require blood sampling at medical institutions, providing a convenient and practical method for predicting hypertensive disorders of pregnancy.
- The negative predictive value for prediction within one week was greater than 98%, suggesting that the proposed method would be especially useful to screen out low risk individuals, contributing to reducing medical consultations.

## Introduction

Hypertensive disorders of pregnancy (HDP) are hypertension and related symptoms in pregnant women. The HDP incidence is approximately 5-10% of pregnant women [1]. HDP is classified into four subtypes: gestational hypertension (GH), preeclampsia (PE), chronic hypertension, and superimposed preeclampsia [2-5]. Among them, GH and PE are predominant and are characterized by new onset of hypertension after 20 weeks of gestation [6]. PE causes more than 70,000 maternal deaths and more than 500,000 fetal and neonatal deaths worldwide each year [3]. Furthermore, approximately 15-25% of GH cases may progress to PE [7]. Therefore, establishing an early prediction method for PE and GH is needed.

Previous studies have developed prediction algorithms for PE based on biophysical markers including mean arterial pressure and uterine artery pulsatility index, biochemical markers including placental growth factor (PlGF), maternal demographic characteristics, and medical history [8-14]. In addition, the ratio of soluble fms-like tyrosine kinase-1 (sFlt-1) to PlGF has demonstrated high predictive efficacy [15,16]. The sFlt-1/PlGF test was approved in Japan in July 2021 and in the United States in May 2023. However, a limitation of existing prediction methods is their reliance on serum markers, such as PlGF and sFlt-1, which require blood sampling.

In contrast, home blood pressure monitoring/measurement (HBPM) enables frequent, accurate, and long-term measurements at low cost without white coat effects [17,18]. HBPM is also expected to be a promising technology for HDP screening [2,19,20]. However, no machine-learning model has been developed so far to predict HDP based on HBPM due to the complexity of blood pressure variations during pregnancy.

To resolve this issue, we focused on the Babies and their Parents’ Longitudinal Observation in Suzuki Memorial Hospital in Intrauterine Period (BOSHI) study [21-26], a large-scale cohort study of HBPM during pregnancy. The study revealed detailed long-term blood pressure variations depending on season and gestational age in pregnant women [21]. For normal blood pressure (NBP) cases, the population means of both systolic blood pressure (SBP) and diastolic blood pressure (DBP) increased in winter and decreased in summer [21]. In addition, they reached their lowest levels at around 20 weeks of gestation and increased thereafter toward delivery, forming U-shaped curves [21].

Based on these findings, this study aimed to develop and validate a noninvasive and accurate HDP prediction model based on HBPM, incorporating compensation for the seasonal and gestational age-dependent variations.

## Materials and Methods

### Data and Code Availability

The data and source codes generated, analyzed, or used in the current study are available from the corresponding author upon reasonable request.

### Diagnostic Criteria for Hypertension

Generally, diagnostic criteria for hypertension differ between office blood pressure (OBP) and home blood pressure (HBP) [18,27,28]. However, there are no widely accepted diagnostic criteria for hypertension in pregnancy regarding HBP [29]. Therefore, OBP criteria were used in this study to define hypertension in pregnancy based on HBP: (1) HBP ≥ 140/90 mmHg was observed twice in a row within 48 hours, or (2) HBP ≥ 160/110 mmHg was observed once. OBP monitoring/measurement (OBPM) at prenatal checkups was used only for reference. Supplementary Figure 1 compares HBP and OBP measured on the same day. When all data were analyzed together, median home SBP and DBP were 2.0 and 2.5 mmHg lower than office SBP and DBP, respectively.

Because PE-related symptoms such as proteinuria could not be diagnosed by the participants themselves, detailed diagnosis by a doctor including HDP subtyping was made at a medical institution.

### Study Participants

In the development cohort, we analyzed the dataset of the BOSHI study [21-26], which recruited pregnant women who reserved gave birth at Suzuki Memorial Hospital in Miyagi Prefecture, Japan, from October 2006 to October 2011. Figure 1 shows the flowchart. The total number of participants was 1292 (NBPs: 1051; HDPs: 241). We excluded cases with multiple pregnancy (n=14). We also excluded cases with insufficient HBPM (n=835) defined by: (1) no data within 7 days before hypertension onset or delivery (n=780), (2) less than 14 measurement days after 18 weeks of gestation (n=35), or (3) shorter than 56 days of measurement period including missing values after 11 weeks of gestation (n=20). Finally, the data of 443 participants (NBPs: 378; HDPs: 65) remained and were randomly divided into training data (NBPs: 339; HDPs: 26) and test data (NBPs: 39; HDPs: 39; characteristics matched between NBPs and HDPs for age, prepregnancy body weight, height, delivery month, measurement number, and measurement rate).

**Figure 1.**
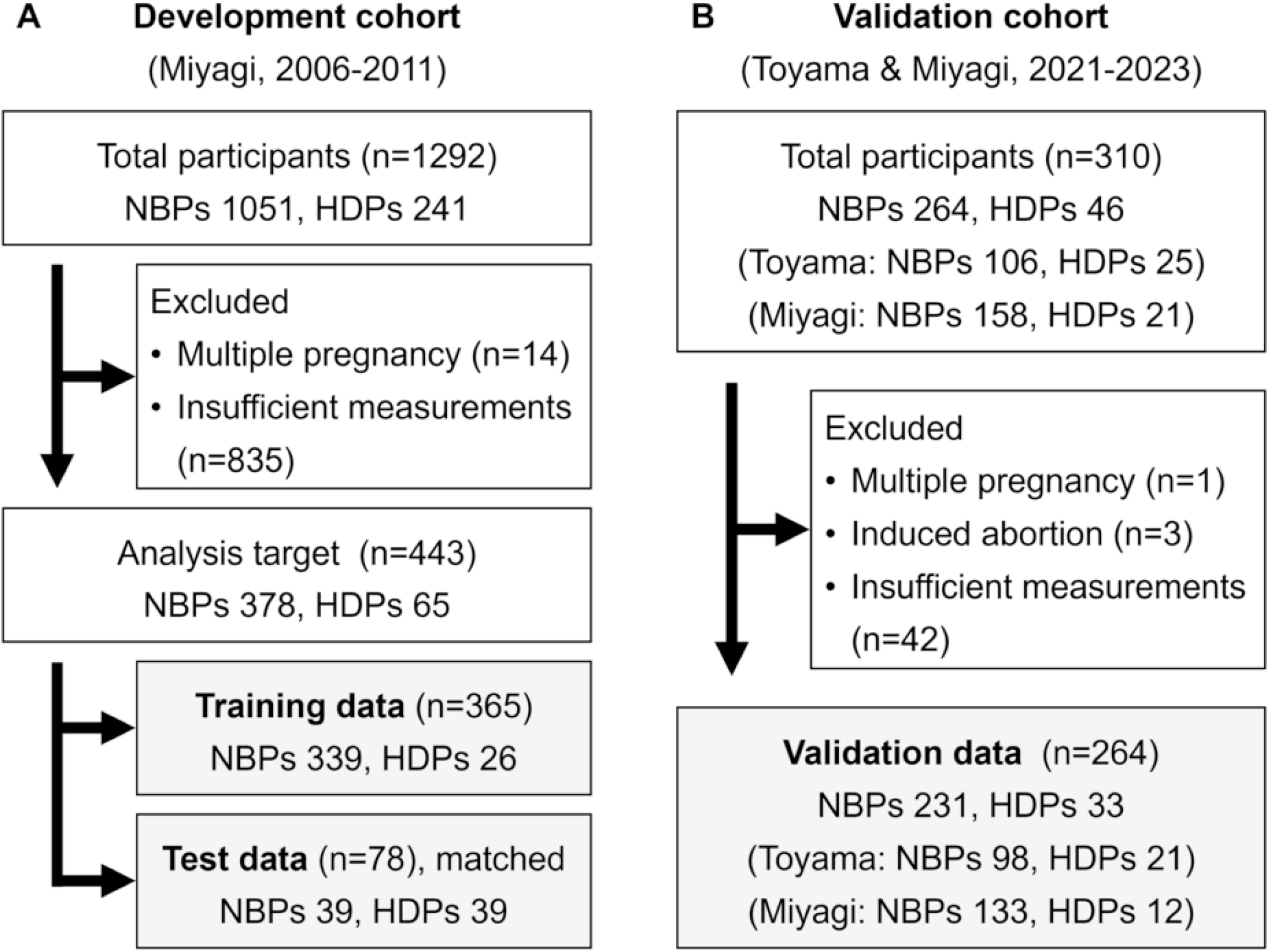
Flow chart. (A) Development cohort. (B) Validation cohort. NBP: normal blood pressure; HDP: hypertensive disorders of pregnancy.

In the validation cohort, HBPM data were prospectively collected from pregnant women who visited either Toyama University Hospital in Toyama Prefecture, Japan, from October 2021 to December 2023, or Suzuki Memorial Hospital from January 2022 to December 2023. We tried to avoid including pregnant women with multiple pregnancy. The total number of participants was 310 (All: NBPs, 264; HDPs, 46; Toyama: NBPs, 106; HDPs, 25; Miyagi: NBPs, 158; HDPs, 21). We excluded cases with multiple pregnancy (n=1) and induced abortion (n=3). We also excluded cases with insufficient HBPM (n=42) defined by: (1) less than 14 measurement days after 18 weeks of gestation (n=35), or (2) shorter than 56 days of measurement period including missing values after 11 weeks of gestation (n=7). Finally, the data of 264 participants (All: NBPs, 231; HDPs, 33; Toyama: NBPs, 98; HDPs, 21; Miyagi: NBPs, 133; HDPs, 12) remained and were used as validation data.

### Data Collection

The development cohort data were extracted from the database of the BOSHI study [21-26]. In the BOSHI study, the participants were instructed to measure HBP once in the morning and once in the evening every day. The validation cohort data were collected using automatic cuff-style blood pressure monitors (HCR-7501T, Omron Healthcare, Japan), provided to each participant to measure her blood pressure every morning and evening at home. The participants were instructed to measure HBP twice per occasion (that is, twice in the morning and twice in the evening) under conditions recommended by the Japanese Society of Hypertension Guidelines for the Management of Hypertension [28]. The measurement data were transmitted to a dedicated app (OMRON connect, Omron Healthcare, Japan) on each participant’s smartphone via Bluetooth, sent to a secure cloud service (OMRON connect cloud, Omron Healthcare, Japan) over the Internet, and retrieved by authorized personnel using a dedicated service (B2B Data Services Platform, Omron Healthcare, Japan).

The measurement period of the validation cohort was from study participation (median: 11.9 weeks of gestation) to at least delivery and up to 12 weeks after delivery for follow-up. The measurements after delivery or HDP onset were excluded from the analysis for both the development and validation cohorts. Therefore, patients with chronic hypertension or superimposed PE were not included. Table 1 shows HBPM statistics. In the validation cohort, the median number of the total measurements per participant was 360.5, and at least one measurement was taken on 86.2% of all days within the measurement period.

**Table 1.** Statistics of HBPM.

| Item | Development cohort (n=443) | Validation cohort (All, n=264) | Validation cohort (Toyama, n=119) | Validation cohort (Miyagi, n=145) |
| --- | --- | --- | --- | --- |
| Number of total measurements | 245.0<br>(143.0-324.5) | 360.5<br>(251.8-555.8) | 341.0<br>(269.5-396.5) | 404.0<br>(229.0-661.0) |
| Number of measured days | 124.0<br>(78.5-161.0) | 170.5<br>(118.8-189.2) | 173.0<br>(123.5-193.0) | 168.0<br>(99.0-186.0) |
| Number of measurements per day<sup>1</sup> | 1.9<br>(1.7-2.3) | 2.3<br>(1.9-3.4) | 2.0<br>(1.8-2.4) | 2.9<br>(2.2-3.7) |
| Gestational age at start of measurement (week) | 13.4<br>(12.1-15.0) | 11.9<br>(11.0-12.9) | 11.7<br>(10.1-13.4) | 11.9<br>(11.3-12.6) |
| Measurement period (day)<sup>2</sup> | 180.0<br>(166.0-191.0) | 186.0<br>(165.8-196.0) | 184.0<br>(165.0-199.0) | 187.0<br>(168.0-195.0) |
| Proportion of measured days | 69.7% | 86.2% | 91.0% | 82.2% |
| Maximum consecutive missing period (day) | 9.0<br>(4.0-16.0) | 3.0<br>(2.0-7.0) | 2.0<br>(1.5-6.0) | 4.0<br>(2.0-9.0) |
| Morning SBP (mmHg)<sup>3</sup> | 105.0<br>(98.0-112.0) | 102.0<br>(96.0-109.5) | 103.0<br>(96.0-110.0) | 102.0<br>(95.0-109.0) |
| Evening SBP (mmHg)<sup>3</sup> | 104.0<br>(98.0-112.0) | 103.0<br>(96.5-109.8) | 103.0<br>(96.5-110.0) | 102.5<br>(96.5-109.0) |
| Morning DBP (mmHg)<sup>3</sup> | 63.0<br>(57.0-69.0) | 67.0<br>(61.0-72.7) | 67.0<br>(62.0-73.0) | 66.5<br>(60.5-72.5) |
| Evening DBP (mmHg)<sup>3</sup> | 61.0<br>(55.0-67.0) | 65.5<br>(60.0-71.5) | 66.0<br>(60.5-72.0) | 65.0<br>(59.5-71.0) |

2 Numerical data are presented as median and interquartile range. SBP: systolic blood pressure; DBP: 3 diastolic blood pressure.
<sup>1</sup>Days without measurement were excluded. <sup>2</sup>Days without measurement were included. <sup>3</sup>Morning SBP and DBP: 5:00 to 9:00; evening SBP and DBP: 21:00 to 24:00.

### Antihypertensive Medication Use

In the training, test, and validation data, no participants took antihypertensive medications at the beginning of HBPM. In addition, our analysis did not include the period of antihypertensive medication use at all. This is because in all cases, the start of antihypertensive medications was later than delivery or HDP onset (Supplementary Table 1).

### Prediction Model

Figure 2 shows steps of the prediction method for HDP. First, HBPM data were acquired. Each time-stamped blood pressure record was processed individually without aggregation. Second, normalization was performed, in which normal blood pressure variations depending on season and gestational age were subtracted. Supplementary Figure 2 shows the seasonal and gestational age-dependent variations estimated using HBPM data from 12 to 40 weeks of gestation from 339 NBPs in the development cohort. Combinations of trigonometric functions were used for the seasonal variations, and quadratic functions were used for the gestational age-dependent variations. The adjusted SBP and DBP were rescaled to have standard deviations close to 1. Third, missing values were imputed using linear interpolation, with random noise added to the interpolated values to mimic realistic fluctuations. Fourth, at each time point, four features were calculated for the last four weeks: mean SBP, mean DBP, correlation coefficient between SBP and DBP, and slope of SBP against day. The four features were selected from 12 candidates (Supplementary Table 2) based on both their contribution to prediction and domain knowledge. Fifth, the feature values were normalized again. Finally, a weighted summation of the normalized feature values, termed the prediction score, was compared with a threshold. The weights were determined by logistic regression, and the threshold was determined to meet 90% specificity (10% false positive rate) in the training data following previous studies [9-11,13,14]. If the prediction score was greater than or equal to the threshold, HDP development after the prediction time point was expected. Among many machine learning techniques, we selected the logistic regression model due to its simplicity and interpretability.

**Figure 2.**
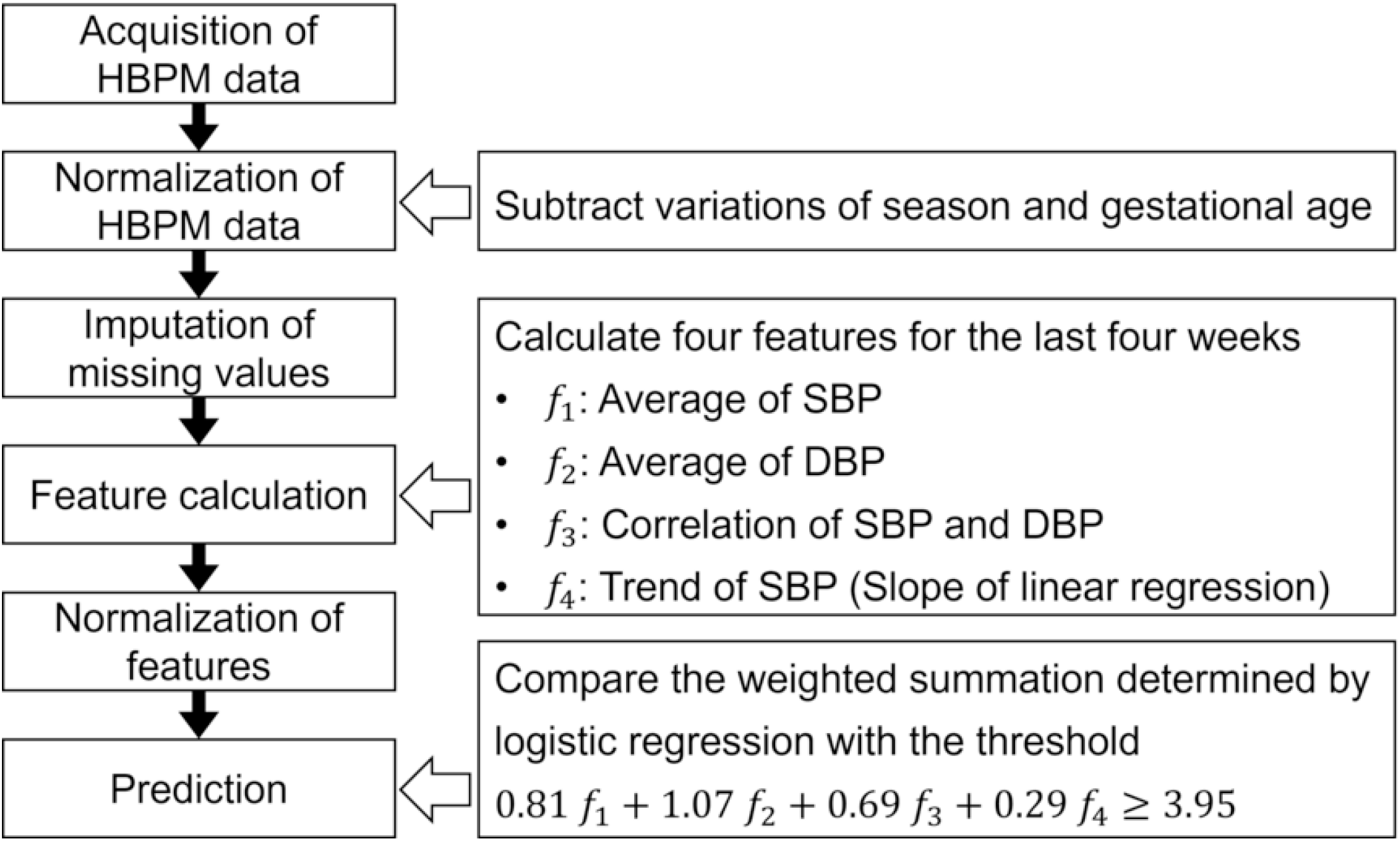
Steps of the prediction method for hypertensive disorders of pregnancy. HBPM: home blood pressure monitoring; SBP: systolic blood pressure; DBP: diastolic blood pressure.

### Statistical Analysis

Statistical analysis was performed with Python version 3.9 (Python Software Foundation, Beaverton, OR, USA). Mann-Whitney *U*-test and chi-squared test were used for between-group comparisons on numerical and categorical variables, respectively. Data were presented as median and interquartile range (IQR). A p-value less than 0.05 was considered statistically significant. The confidence level for confidence intervals (CI) was 95%.

### Ethical Approval

All procedures of this study were performed in accordance with the 1964 Helsinki Declaration, its later amendments, and the “Ethical Guidelines for Life Science and Medical Research Involving Human Subjects” published by the Ministry of Health, Labour and Welfare of Japan. The Institutional Review Boards of Tohoku University School of Medicine, Tohoku Medical and Pharmaceutical University, and the Hospital Review Board of Suzuki Memorial Hospital approved the BOSHI study protocol. The ethics committee of Toyama University Hospital approved the study protocol with respect to Toyama University Hospital on 2021/4/22 (approval number: R2021021), and the ethics committee of Tohoku Medical and Pharmaceutical University approved the study protocol with respect to Suzuki Memorial Hospital on 2022/1/13 (approval number: 2021-0-015-0001). The study participants gave written informed consent.

## Results

The baseline characteristics of the participants are shown in Table 2. Overall, the development and validation cohorts exhibited comparable characteristics. The proportion of prepregnancy obesity (defined as BMI ≥ 25 kg/m^2^ according to the Japanese criterion) was significantly higher in the HDPs compared to the NBPs, both in the development cohort (NBPs: 11.9% (n=45); HDPs: 23.1% (n=15); p=0.025) and the validation cohort (NBPs: 19.5% (n=45); HDPs: 39.4 % (n=13); p=0. 018). Similarly, the median SBP value from the first HBPM was significantly higher in the HDPs compared to the NBPs, both in the development cohort (NBPs: 104.0 (IQR: 98.0-110.0) mmHg; HDPs: 111.0 (IQR: 106.0-117.0) mmHg; p=2.3×10^−9^) and the validation cohort (NBPs: 99.0 (IQR: 92.0-106.0) mmHg; HDPs: 107.0 (IQR: 99.0-113.0) mmHg; p=0.0010). Furthermore, the median DBP value from the first HBPM was also significantly higher in the HDPs compared to the NBPs, both in the development cohort (NBPs: 63.0 (IQR: 58.0-68.0) mmHg; HDPs: 70.0 (IQR: 65.0-75.0) mmHg; p=3.1×10^−9^) and the validation cohort (NBPs: 67.0 (IQR: 62.0-72.0) mmHg; HDPs: 75.0 (IQR: 71.0-79.0) mmHg; p=3.0×10^−5^).

**Table 2.** Baseline characteristics of the participants.

| Item | Development<br>cohort<br>(NBPs) | Development<br>cohort<br>(HDPs) | Development<br>cohort<br>(p-value) | Validation<br>cohort<br>(NBPs) | Validation<br>cohort<br>(HDPs) | Validation<br>cohort<br>(p-value) |
| --- | --- | --- | --- | --- | --- | --- |
| Participants (n) | 378 (85.3%) | 65 (14.7%) | - | 231 (87.5%) | 33 (12.5%) | - |
| Age (year) | 32.0<br>(28.0-35.0) | 34.0<br>(29.0-36.0) | 0.051 | 33.0<br>(29.0-37.0) | 34.0<br>(32.0-38.0) | 0.074 |
| Prepregnancy BMI<br>(kg/m<sup>2</sup>) | 20.6<br>(19.4-22.8) | 21.8<br>(19.8-24.5) | 0.028 | 21.6<br>(19.5-24.2) | 23.4<br>(20.7-28.3) | 0.029 |
| Prepregnancy obesity<br>(n) | 45 (11.9%) | 15 (23.1%) | 0.025 | 45 (19.5%) | 13 (39.4%) | 0.018 |
| SBP (mmHg)<sup>1</sup> | 104.0<br>(98.0-110.0) | 111.0<br>(106.0-117.0) | 2.3×10<sup>-9</sup> | 99.0<br>(92.0-106.0) | 107.0<br>(99.0-113.0) | 0.0010 |
| DBP (mmHg)<sup>1</sup> | 63.0<br>(58.0-68.0) | 70.0<br>(65.0-75.0) | 3.1×10<sup>-9</sup> | 67.0<br>(62.0-72.0) | 75.0<br>(71.0-79.0) | 3.0×10<sup>-5</sup> |
| Smoking, past (n) | 55 (14.6%) | 9 (13.8%) | 0.97 | 25 (10.8%) | 1 (3.0%) | 0.27 |
| Smoking, current (n) | 12 (3.2%) | 2 (3.1%) | 0.73 | 1 (0.4%) | 0 (0.0%) | 0.26 |
| Nullipara (n) | 160 (42.3%) | 27 (41.5%) | 0.99 | 69 (29.9%) | 11 (33.3%) | 0.84 |
| Multipara with past<br>HDP history (n) | 8 (2.1%) | 4 (6.2%) | 0.15 | 15 (6.5%) | 7 (21.2%) | 0.012 |
| Parent history of<br>hypertension (n) | 141 (37.3%) | 31 (47.7%) | 0.15 | 84 (36.4%) | 14 (42.4%) | 0.63 |
| Parent history of<br>diabetes (n) | 63 (16.7%) | 8 (12.3%) | 0.48 | 31 (13.4%) | 3 (9.1%) | 0.68 |

2 Numerical data are presented as median and interquartile range. Prepregnancy obesity: prepregnancy BMI 3 (body mass index) $\geq 25$ kg/m<sup>2</sup>. NBP: normal blood pressure; HDP: hypertensive disorders of 4 pregnancy; SBP: systolic blood pressure; DBP: diastolic blood pressure.
<sup>1</sup>The SBP and DBP values shown are from the first home blood pressure measurement.

The adverse pregnancy outcomes of the participants are shown in Table 3. The median gestational age at delivery was significantly lower in the HDPs compared to the NBPs in the development cohort (NBPs: 39.9 (IQR: 39.0-40.6) weeks; HDPs: 39.4 (IQR: 38.4-40.1) weeks; p=0.0021). The median of mean SBP during the third trimester was significantly higher in the HDPs compared to the NBPs, both in the development cohort (NBPs: 105.0 (IQR: 100.1-109.8) mmHg; HDPs: 119.3 (IQR: 113.3-123.8) mmHg; p=9.3×10^−24^) and the validation cohort (NBPs: 104.7 (IQR: 100.2-110.5) mmHg; HDPs: 115.3 (IQR: 110.4-119.3) mmHg; p=3.9×10^−10^). Similarly, the median of mean DBP during the third trimester was significantly higher in the HDPs compared to the NBPs, both in the development cohort (NBPs: 62.0 (IQR: 58.4-65.9) mmHg; HDPs: 75.1 (IQR: 70.3-77.9) mmHg; p=1.9×10^−26^) and the validation cohort (NBPs: 67.1 (IQR: 62.6-71.8) mmHg; HDPs: 78.3 (IQR: 74.6-81.0) mmHg; p=3.3×10^−12^).

**Table 3.** Adverse pregnancy outcomes of the participants.

| Item | Development cohort<br>(NBPs, n=378) | Development cohort<br>(HDPs, n=65) | Development cohort<br>(p-value) | Validation cohort<br>(NBPs, n=231) | Validation cohort<br>(HDPs, n=33) | Validation cohort<br>(p-value) |
| --- | --- | --- | --- | --- | --- | --- |
| Gestational age at delivery (week) | 39.9<br>(39.0-40.6) | 39.4<br>(38.4-40.1) | 0.0021 | 39.2<br>(38.3-40.1) | 39.1<br>(38.4-40.3) | 0.82 |
| Preterm delivery (n) | 7 (1.9%) | 4 (6.2%) | 0.10 | 12 (5.2%) | 0 (0.0%) | 0.37 |
| Mean SBP during 3rd trimester (mmHg) | 105.0<br>(100.1-109.8) | 119.3<br>(113.3-123.8) | $9.3 \times 10^{-24}$ | 104.7<br>(100.2-110.5) | 115.3<br>(110.4-119.3) | $3.9 \times 10^{-10}$ |
| Mean DBP during 3rd trimester (mmHg) | 62.0<br>(58.4-65.9) | 75.1<br>(70.3-77.9) | $1.9 \times 10^{-26}$ | 67.1<br>(62.6-71.8) | 78.3<br>(74.6-81.0) | $3.3 \times 10^{-12}$ |
| Max SBP during 3rd trimester (mmHg) | 122.0<br>(114.0-129.0) | 144.0<br>(134.0-154.0) | $3.1 \times 10^{-25}$ | 127.0<br>(119.0-134.2) | 147.0<br>(138.0-156.0) | $7.1 \times 10^{-13}$ |
| Max DBP during 3rd trimester (mmHg) | 77.0<br>(72.0-82.0) | 95.0<br>(89.0-100.0) | $8.4 \times 10^{-26}$ | 84.0<br>(77.0-92.0) | 102.0<br>(98.0-106.0) | $1.3 \times 10^{-15}$ |
| GDM (n) | n/a | n/a | - | 19 (8.2%) | 2 (6.1%) | 0.93 |
| Proteinuria (n) | 7 (1.9%) | 3 (4.6%) | 0.35 | 2 (0.9%) | 1 (3.0%) | 0.83 |
| FGR (n) | 10 (2.6%) | 2 (3.1%) | 0.83 | 17 (7.4%) | 3 (9.1%) | 1.0 |
| NRFS (n) | 27 (7.1%) | 12 (18.5%) | 0.0062 | 20 (8.7%) | 3 (9.1%) | 0.80 |
| Caesarian section (n) | 54 (14.3%) | 29 (44.6%) | $1.9 \times 10^{-8}$ | 76 (32.9%) | 9 (27.3%) | 0.65 |
| birth weight (g) | 3074<br>(2816-3318) | 2978<br>(2728-3180) | 0.027 | 3036<br>(2854-3272) | 3031<br>(2796-3415) | 0.76 |
| LBW (n) | 21 (5.6%) | 9 (13.8%) | 0.029 | 20 (8.7%) | 3 (9.1%) | 0.80 |
| SGA (n) | 36 (9.5%) | 4 (6.2%) | 0.52 | 13 (5.6%) | 3 (9.1%) | 0.70 |
| Macrosomia (n) | 1 (0.3%) | 0 (0.0%) | 0.32 | 0 (0.0%) | 0 (0.0%) | - |
| LGA (n) | 37 (9.8%) | 2 (3.1%) | 0.13 | 15 (6.5%) | 4 (12.1%) | 0.42 |
| Apgar score at 5 min < 7 (n) | 0 (0.0%) | 0 (0.0%) | - | 1 (0.4%) | 0 (0.0%) | 0.26 |

2 Numerical data are presented as median and interquartile range. Preterm delivery: delivery before 37 3 weeks. LBW (low birth weight): birth weight < 2500 g. Macrosomia: birth weight ≥ 4000 g. SGA 4 (small for gestational age): birth weight below the 10th percentile for gestational age. LGA (large for 5 gestational age): birth weight above the 90th percentile for gestational age. NBP: normal blood 6 pressure; HDP: hypertensive disorders of pregnancy; SBP: systolic blood pressure; DBP: diastolic 7 blood pressure; GDM: gestational diabetes mellitus; FGR: fetal growth restriction; NRFS: non- 8 reassuring fetal status.

Figure 3 shows the receiver operating characteristic (ROC) curves of the training, test, and validation data. The area under the curve (AUC) values were 0.949 (CI: 0.890-1.000), 0.884 (CI: 0.807-0.961), and 0.845 (CI: 0.759-0.930) for the training, test, and validation data, respectively. In other words, our prediction method consistently showed high AUC values.

**Figure 3.**
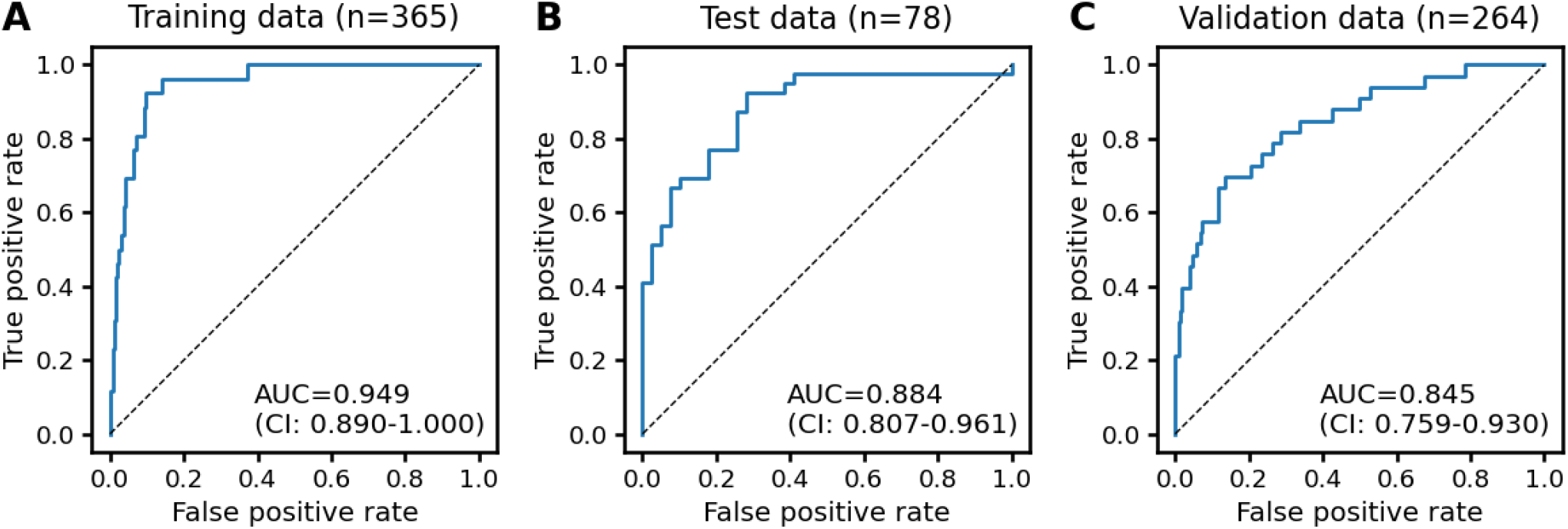
Receiver operating characteristic curves. (A) Training data. (B) Test data. (C) Validation data. AUC: area under the curve; CI: confidence interval.

Table 4 shows the prediction performance of the proposed method. For the training data, sensitivity, specificity, positive predictive value (PPV), and negative predictive value (NPV) were 0.923 (CI: 0.749-0.991), 0.888 (CI: 0.849-0.919), 0.387 (CI: 0.266-0.519), and 0.993 (CI: 0.976-0.999), respectively. For the test data, sensitivity, specificity, PPV, and NPV were 0.667 (CI: 0.498-0.809), 0.897 (CI: 0.758-0.971), 0.867 (CI: 0.693-0.962), and 0.729 (CI: 0.582-0.847), respectively. Note that some statistics depend on the balance between the NBPs and HDPs, and the proportion of the HDPs in the test data was 50% due to matching, which was much higher than the original population. For the validation data, sensitivity, specificity, PPV, and NPV were 0.758 (CI: 0.577-0.889), 0.766 (CI: 0.706-0.819), 0.316 (CI: 0.216-0.431), and 0.957 (CI: 0.917-0.981), respectively. These results confirmed that our prediction method could consistently exhibit high NPV as well as moderate sensitivity and specificity.

**Table 4.** Prediction performance of the proposed method.

|  | Sensitivity | Specificity | PPV | NPV | PLR | NLR |
| --- | --- | --- | --- | --- | --- | --- |
| Development cohort,<br>training (n=365;<br>NBPs: 339; HDPs: 26) | 0.923<br>(0.749-0.991) | 0.888<br>(0.849-0.919) | 0.387<br>(0.266-0.519) | 0.993<br>(0.976-0.999) | 8.235<br>(5.983-11.33) | 0.087<br>(0.023-0.328) |
| Development cohort,<br>test (n=78; NBPs: 39;<br>HDPs: 39) | 0.667<br>(0.498-0.809) | 0.897<br>(0.758-0.971) | 0.867<br>(0.693-0.962) | 0.729<br>(0.582-0.847) | 6.500<br>(2.502-16.88) | 0.371<br>(0.235-0.586) |
| Validation cohort,<br>Toyama (n=119;<br>NBPs: 98; HDPs: 21) | 0.762<br>(0.528-0.918) | 0.776<br>(0.680-0.854) | 0.421<br>(0.263-0.592) | 0.938<br>(0.862-0.980) | 3.394<br>(2.188-5.264) | 0.307<br>(0.142-0.665) |
| Validation cohort,<br>Miyagi (n=145;<br>NBPs: 133; HDPs: 12) | 0.750<br>(0.428-0.945) | 0.759<br>(0.678-0.829) | 0.220<br>(0.106-0.376) | 0.971<br>(0.918-0.994) | 3.117<br>(1.998-4.863) | 0.329<br>(0.123-0.881) |
| Validation cohort,<br>all (n=264; NBPs:<br>231; HDPs: 33) | 0.758<br>(0.577-0.889) | 0.766<br>(0.706-0.819) | 0.316<br>(0.216-0.431) | 0.957<br>(0.917-0.981) | 3.241<br>(2.394-4.387) | 0.316<br>(0.172-0.581) |

2 The 95% confidence intervals are in parentheses. PPV: positive predictive value; NPV: negative 3 predictive value; PLR: positive likelihood ratio; NLR: negative likelihood ratio.

As a sensitivity analysis, we investigated how the prediction performance changes if hypertension was redefined using HBP criteria for the general population: (1) HBP ≥ 135/85 mmHg was observed twice in a row within 48 hours, or (2) HBP ≥ 155/105 mmHg was observed once. Supplementary Table 3 shows prediction performance of the proposed method in the sensitivity analysis. The results did not change much, indicating the robustness of our prediction method.

As a control, we investigated prediction performance of a baseline model that uses only prepregnancy BMI, SBP from the first HBPM, and DBP from the first HBPM as explanatory variables. For the baseline model, logistic regression was used, and the threshold was determined to meet 90% specificity in the training data, similarly to the proposed model. Supplementary Table 4 shows the prediction performance of the baseline model. Its overall performance was worse than the proposed model. Especially, its sensitivity was 0.182 for the validation data, which was much lower than that of the proposed model. In other words, the high prediction performance of the proposed model could not be achieved by the simple baseline model.

Table 5 shows the prediction performance of the proposed method for predictions within one week or four weeks. Because the sFlt-1/PlGF ratio is used for predicting the absence of HDP within one week and the occurrence of HDP within four weeks, we focused on NPV within one week and PPV within four weeks. For the training data, NPV within one week was 0.999 (CI: 0.998-0.999), and PPV within four weeks was 0.299 (CI: 0.282-0.317). For the test data, NPV within one week was 0.983 (CI: 0.981-0.985), and PPV within four weeks was 0.407 (CI: 0.381-0.432). For the validation data, NPV within one week was 0.998 (CI: 0.997-0.998), and PPV within four weeks was 0.168 (CI: 0.154-0.182). These results confirmed that our prediction method consistently showed quite high NPV within one week.

**Table 5.** Prediction performance of the proposed method for predictions within one week or four weeks.

|  | NPV<br>(within 1 w) | Sensitivity<br>(within 1 w) | Specificity<br>(within 1 w) | PPV<br>(within 4 w) | Sensitivity<br>(within 4 w) | Specificity<br>(within 4 w) |
| --- | --- | --- | --- | --- | --- | --- |
| Development cohort,<br>training (n=365;<br>NBPs: 339; HDPs: 26) | 0.999<br>(0.998-0.999) | 0.733<br>(0.687-0.776) | 0.970<br>(0.969-0.972) | 0.299<br>(0.282-0.317) | 0.657<br>(0.629-0.684) | 0.976<br>(0.975-0.977) |
| Development cohort,<br>test (n=78; NBPs: 39;<br>HDPs: 39) | 0.983<br>(0.981-0.985) | 0.470<br>(0.424-0.517) | 0.918<br>(0.914-0.923) | 0.407<br>(0.381-0.432) | 0.341<br>(0.318-0.363) | 0.938<br>(0.934-0.942) |
| Validation cohort,<br>Toyama (n=119;<br>NBPs: 98; HDPs: 21) | 0.997<br>(0.996-0.998) | 0.657<br>(0.571-0.736) | 0.902<br>(0.897-0.906) | 0.184<br>(0.165-0.203) | 0.543<br>(0.501-0.585) | 0.913<br>(0.908-0.917) |
| Validation cohort,<br>Miyagi (n=145;<br>NBPs: 133; HDPs: 12) | 0.999<br>(0.998-0.999) | 0.733<br>(0.619-0.829) | 0.927<br>(0.923-0.931) | 0.148<br>(0.129-0.168) | 0.639<br>(0.582-0.693) | 0.935<br>(0.931-0.938) |
| Validation cohort, all<br>(n=264; NBPs: 231;<br>HDPs: 33) | 0.998<br>(0.997-0.998) | 0.684<br>(0.617-0.746) | 0.915<br>(0.912-0.918) | 0.168<br>(0.154-0.182) | 0.577<br>(0.543-0.610) | 0.924<br>(0.921-0.927) |

2 The 95% confidence intervals are in parentheses. NPV: negative predictive value; PPV: positive 3 predictive value.

Supplementary Figure 3 shows ROC curves for predictions within one week or four weeks. AUC within one week were 0.963 (CI: 0.950-0.976), 0.834 (CI: 0.810-0.857), and 0.879 (CI: 0.849-0.909) for the training, test, and validation data, respectively. AUC within four weeks were 0.933 (CI: 0.923-0.943), 0.799 (CI: 0.787-0.812), and 0.844 (CI: 0.828-0.861) for the training, test, and validation data, respectively. These results confirmed that our method’s ROC-AUC values were also high for predictions within one week or four weeks.

## Discussion

In this study, we developed an HDP prediction model based on HBPM in the development cohort and verified its effectiveness in the validation cohort. Our method demonstrated high NPV and moderate sensitivity and specificity across the training, test, and validation data. The high NPV suggests suitability for screening individuals at low risk for HDP, reducing unnecessary medical tests or interventions. Another advantage of our method is that it does not require blood sampling, as is the case with the existing prediction methods [8-16].

Notably, our method has a sufficiently high NPV within one week (training data: 0.999; test data: 0.983; validation data: 0.998), which is comparable to that of the sFlt-1/PlGF ratio [15] (its development cohort: 0.989; its validation cohort: 0.993). However, PPV within four weeks of our method (training data: 0.299; test data: 0.407; validation data: 0.168) was, except for the case of test data, lower than that of the sFlt-1/PlGF ratio [15] (its development cohort: 0.407; its validation cohort: 0.367). The difference is that our cases involved healthy pregnant women without risk factors, whereas Zeisler et al. [15] studied cases with clinical suspicion of preeclampsia. In fact, ROC-AUC within one week was comparable between our method (training data: 0.963; test data: 0.834; validation data: 0.879) and that of the sFlt-1/PlGF ratio [15] (its development cohort: 0.898; its validation cohort: 0.861). Similarly, ROC-AUC within four weeks was not quite different between our method (training data: 0.933; test data: 0.799; validation data: 0.844) and the sFlt-1/PlGF ratio [15] (its development cohort: 0.861; its validation cohort: 0.823).

One of the strengths of our study is the use of large-scale and high-quality cohort data from the BOSHI study [21-26], which enabled accurate modeling of seasonal and gestational age-dependent blood pressure variations for reliable predictions. Another strength is the validation of our method in a separate multicenter cohort. It has been reported that models developed and tested within the same cohort often exhibit high predictive performance, but their accuracy frequently declines when applied to temporally or geographically distinct external cohorts due to overfitting and limited generalizability [30,31]. Our study addresses this by explicitly quantifying the drop in predictive performance and demonstrating the feasibility and limitations of model transferability across populations.

Moreover, the use of HBPM itself has many advantages [17,18] and has been expected to be useful for HDP prediction [2,19,20]. Hsieh et al. used a machine learning model based on HBPM to predict cardiovascular outcomes in the general adult population [32]. Although previous studies reported the clinical value of HBPM in pregnant women at high risk of HDP [33,34], to our knowledge, this study is the first to demonstrate the effectiveness of HBPM in predicting HDP in the general pregnant population. A randomized controlled trial demonstrated that HBPM can help reduce prenatal checkups without increasing maternal anxiety [35]. Our method may contribute to safely omitting prenatal HDP screening at medical institutions for low-risk pregnant women.

Our study also confirmed the feasibility of HBPM in pregnant women. In our study, the participants in the validation cohort measured blood pressure on 86.2% of the days within the approximately six-month measurement period (Table 1). The good adherence was probably due to the ease of HBPM. Interestingly, there was a slight improvement in measurement frequency in the validation cohort compared to the development cohort. A possible reason could be that, in the validation cohort, we used a commercially available blood pressure monitor integrated with a dedicated smartphone app provided by the manufacturer. The smartphone app was useful for the participants to check their blood pressure history and to show it to clinical staff during prenatal checkups, which may have helped to maintain motivation. The accuracy of HBPM and the reduction of white coat effects have also been confirmed (Supplementary Figure 1).

The threshold was determined to meet 90% specificity according to previous studies [9-11,13,14]. Given that our model proved to be particularly suitable for screening low risk individuals, adjusting the threshold for that purpose may add more clinical value. However, NPV within one week was already quite high, exceeding 99% in the validation data. Therefore, the current threshold would be practical for rule-out purposes, given that the prediction score is updated each time HBP is measured.

Although detection of HDP after its onset by HBPM seems sufficient, early prediction of HDP has additional advantages. First, HDP prediction may reduce medical consultations for low-risk individuals. Second, HDP prediction allows for targeted proactive health care for high-risk individuals, such as lifestyle modifications and drug treatments under medical supervision, which may reduce the incidence and severity of HDP. Third, HDP prediction may improve adherence to HBPM in individuals predicted to develop HDP in the near future, reducing missed measurements. In fact, HBPM was not performed on 13.8% of days in our validation cohort, and missing values sometimes lasted for several days. Therefore, early prediction may also increase the detection rate of HDP through promotion of HBPM.

It is generally expected that HBPM can detect increases in blood pressure earlier than OBPM. A previous study reported that the first episode of HBP ≥ 140/90 mmHg occurred approximately one month (median: 29 days) before clinical hypertension diagnosis [36]. However, our data showed no significant difference between the time when HBP and OBP first exceeded 140/90 mmHg (development cohort: median difference, 0 day; IQR, −8.5 to 5 days, p=0.67, Wilcoxon test; validation cohort: median difference, 1 day; IQR, −3 to 19 days, p=0.50, Wilcoxon test). This discrepancy may be due to white coat effects; some participants had a consistent increase of 10 mmHg or more in OBP compared with HBP. In addition, the TMM BirThree Cohort Study showed that both HBP and research BP were significantly lower than OBP [37], which may have influenced the timing of hypertension detection in our setting.

Our study has some limitations. First, this study is an observational study, not an intervention study. It is an important future work to evaluate the effectiveness of early interventions based on our prediction model, such as lifestyle modifications and drug treatments under medical supervision. Second, our study was based only on pregnant women in Toyama and Miyagi prefectures in Japan, limiting the generalizability of our findings. Future studies on more diverse populations are needed to expand the applicability of our model. Given that the ROC-AUC value slightly decreased from 0.884 for the test data to 0.845 for the validation data, the predictive performance of our model may further decrease when applied to other populations. If our model is applied to populations largely different from those of the present study, such as non-Japanese people, it would be reasonable to recalculate the seasonal and gestational age-dependent blood pressure variations. For example, in countries with small annual temperature variations, there is little need to correct the seasonal blood pressure variation. The optimal parameters for the adjustment should be determined according to the climate and residential environment of each country. Indeed, an international study has demonstrated substantial heterogeneity in the health effects of ambient temperature across countries, with particularly pronounced impacts observed in populations living in mid-latitude regions where both cold and heat extremes contribute significantly to health risks [38]. This suggests that the seasonal correction derived from our Japanese cohort, which itself lies in a mid-latitude climate, may not be directly transferable to populations in different climatic zones, thereby warranting population-specific recalibration when applying our model to non-Japanese settings. Third, our prediction method has not been fully automated. Integrating our model into existing devices, software, and platforms, such as blood pressure monitors, smart phone apps, and cloud services, could facilitate the use of our model. Fourth, the HDP proportion in the test data was 50% due to individual-level matching, and thus the PPV, NPV, PLR, and NLR of the test data should be interpreted with caution. Fifth, a direct comparison of the proposed method with the sFlt-1/PlGF ratio was missing. It is an important future work to perform both the proposed method and the sFlt-1/PlGF ratio test to the same individuals. Finally, our model has low PPV within four weeks in our validation cohort. To refine the prediction model, parameter optimization using larger data is needed. In addition, incorporating maternal demographic characteristics and medical history could potentially improve the predictive accuracy of the model.

## Conclusions

In conclusion, we developed an HDP prediction model based on HBPM and verified its effectiveness. Especially, our method consistently exhibited high NPV values, and thus it would be useful to screen out normotensive individuals, which may contribute to reducing medical consultations. Further studies are needed to determine whether it is effective to start interventions in pregnant women at the time when the HDP onset is predicted by the prediction model. In addition, integrating the prediction model into blood pressure monitors, smart phone apps, and cloud services is another important future work. We hope that HBPM will become the standard for pregnant women in the near future, and that accurate prediction of HDP based on HBPM and appropriate early interventions will save both maternal and neonatal health.

## Acknowledgements

We would like to thank the study participants and clinical staff for their efforts in data collection.

## Sources of Funding

This research was supported by JST Moonshot R&D Grant Number JPMJMS2021 and JSPS KAKENHI Grant Number JP25K02891. The BOSHI study was supported by MEXT/JSPS KAKENHI Grant Numbers JP16H05243, JP17K15857, JP18390192, JP18590587, JP18K15837, JP19H03905, JP19K18659, JP21390201, JP21K10438, JP24689061, JP25253059, JP25K02891, and JP26860412; MHLW Program Grant Number JPMH19DA1001 and H21-Junkankitou [Seishuu]-Ippan-004; Children and Families Agency Program Grant Numbers JPCA23DA0601 and JPCA24DA0101; AMED Grant Numbers JP19gk0110039 and JP24gn0110088; a Grant-in-Aid for JSPS Fellows (19.7152).

## Disclosures

For all authors, no potential conflicts of interest relevant to this study were reported. However, the following may represent potential conflicts of interest: HM received academic contributions from the J&J Medical Research Grant; HM received scholarship donations from Chugai Pharmaceutical Co., Ltd., Daiichi Sankyo Co., Ltd., and Otsuka Pharmaceutical Co., Ltd.; HM concurrently holds a non-compensated sub-directorship position at the Tohoku Institute for Management of Blood Pressure, supported by Omron Health Care Co. Ltd., and is involved in collaborative research with Omron Health Care in another study.

## Author contributions

Conceptualization: HM, SS. Methodology: SO. Software: MO, SO. Formal Analysis: MO, SO. Investigation: HM, EA, MI, AS, AN, RT, SY, NY, STs, STa, KH, KT, MY. Data Curation: MO, SO, HM, EA, MI, RT. Writing – Original Draft: MO. Writing – Review & Editing: All authors. Supervision: SS. Funding acquisition: HM, SS.

## Abbreviations and Acronyms

AUC: Area Under the Curve
BMI: Body Mass Index
BOSHI study: Babies and Their Parents’ Longitudinal Observation in Suzuki Memorial Hospital in Intrauterine Period study
CI: Confidence Interval
DBP: Diastolic Blood Pressure
GH: Gestational Hypertension
HBP: Home Blood Pressure
HBPM: Home Blood Pressure Monitoring/Measurement
HDP: Hypertensive Disorders of Pregnancy
IQR: Interquartile Range
NBP: Normal Blood Pressure
NPV: Negative Predictive Value
OBP: Office Blood Pressure
OBPM: Office Blood Pressure Monitoring/Measurement
PE: Preeclampsia
PlGF: Placental Growth Factor
PPV: Positive Predictive Value
ROC: Receiver Operating Characteristic
SBP: Systolic Blood Pressure
sFlt-1: Soluble Fms-like Tyrosine Kinase-1

## Notes

### Author Declarations

The ethics committee of Toyama University Hospital gave ethical approval for this work with respect to Toyama University Hospital on 2021/4/22 (approval number: R2021021), and the ethics committee of Tohoku Medical and Pharmaceutical University gave ethical approval for this work with respect to Suzuki Memorial Hospital on 2022/1/13 (approval number: 2021-0-015-0001).

### Summary of Updates

Updated title, abstract, main text, references, figures, tables, and supplementary material.

