## Supplemental Material for "Prediction model of hypertensive disorders of pregnancy based on home blood pressure monitoring"

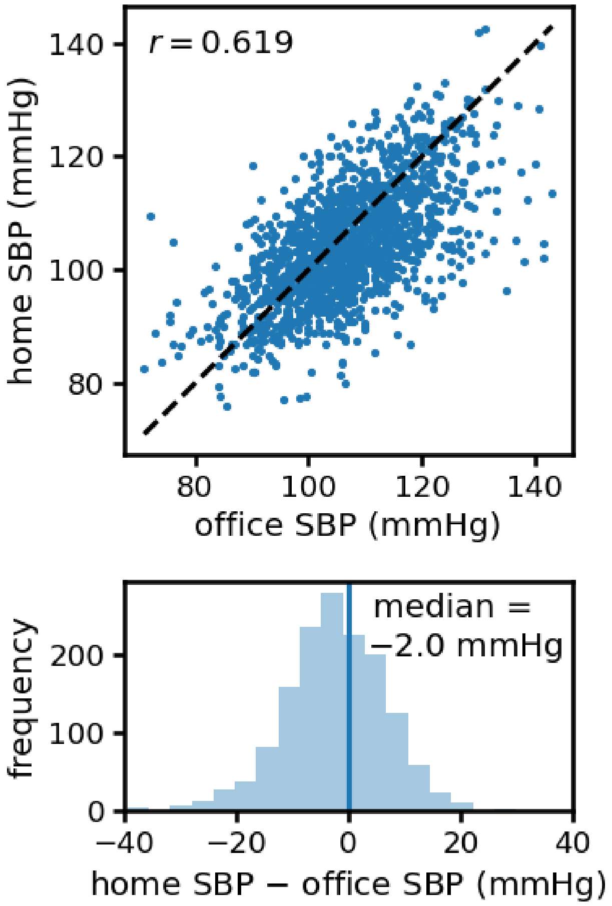


Supplementary Figure 1: Comparison of home blood pressure and office blood pressure measured on the same day in the validation cohort.

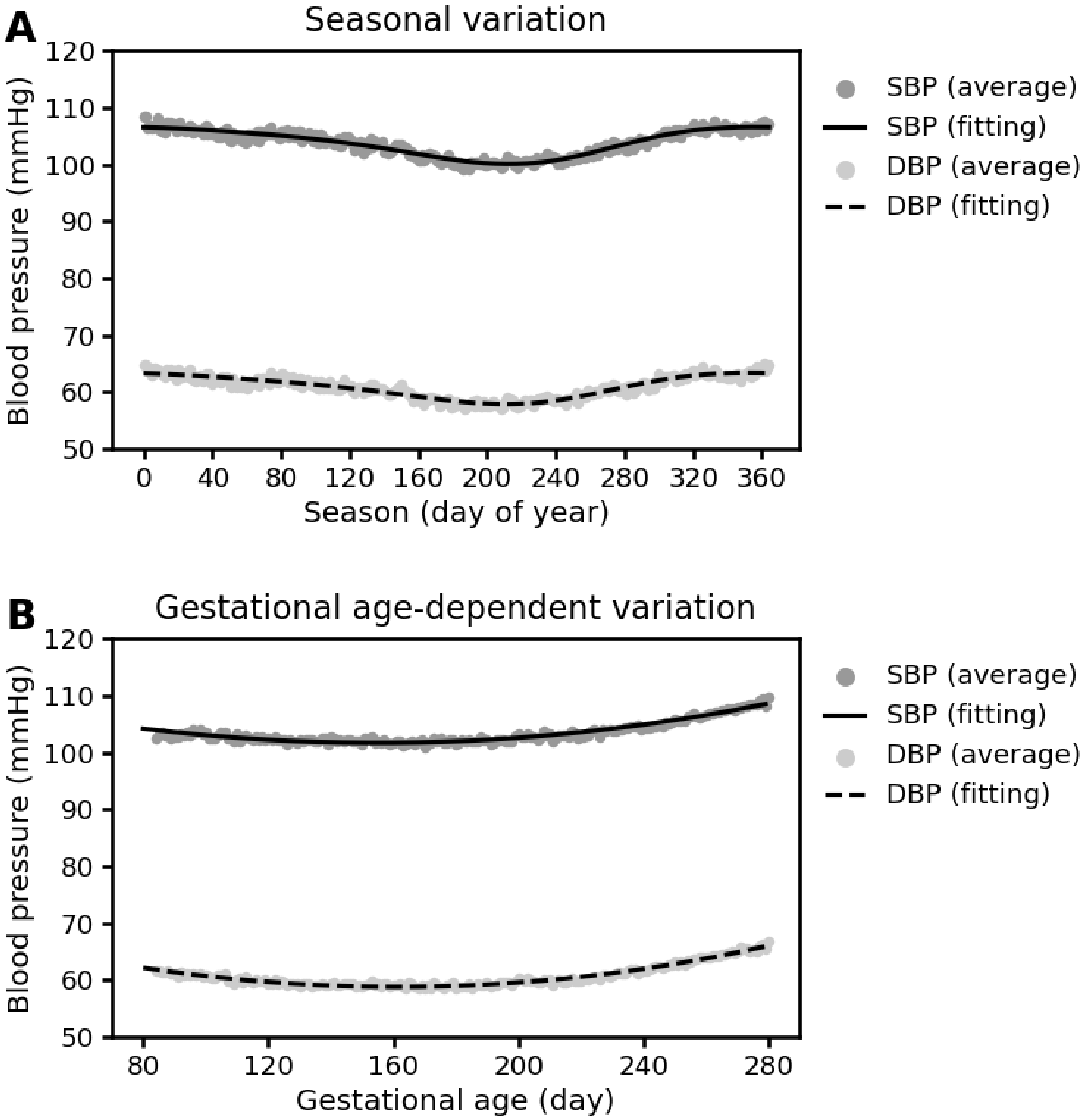


Supplementary Figure 2. Blood pressure variations. (A) Seasonal variation. (B) Gestational age-dependent variation.

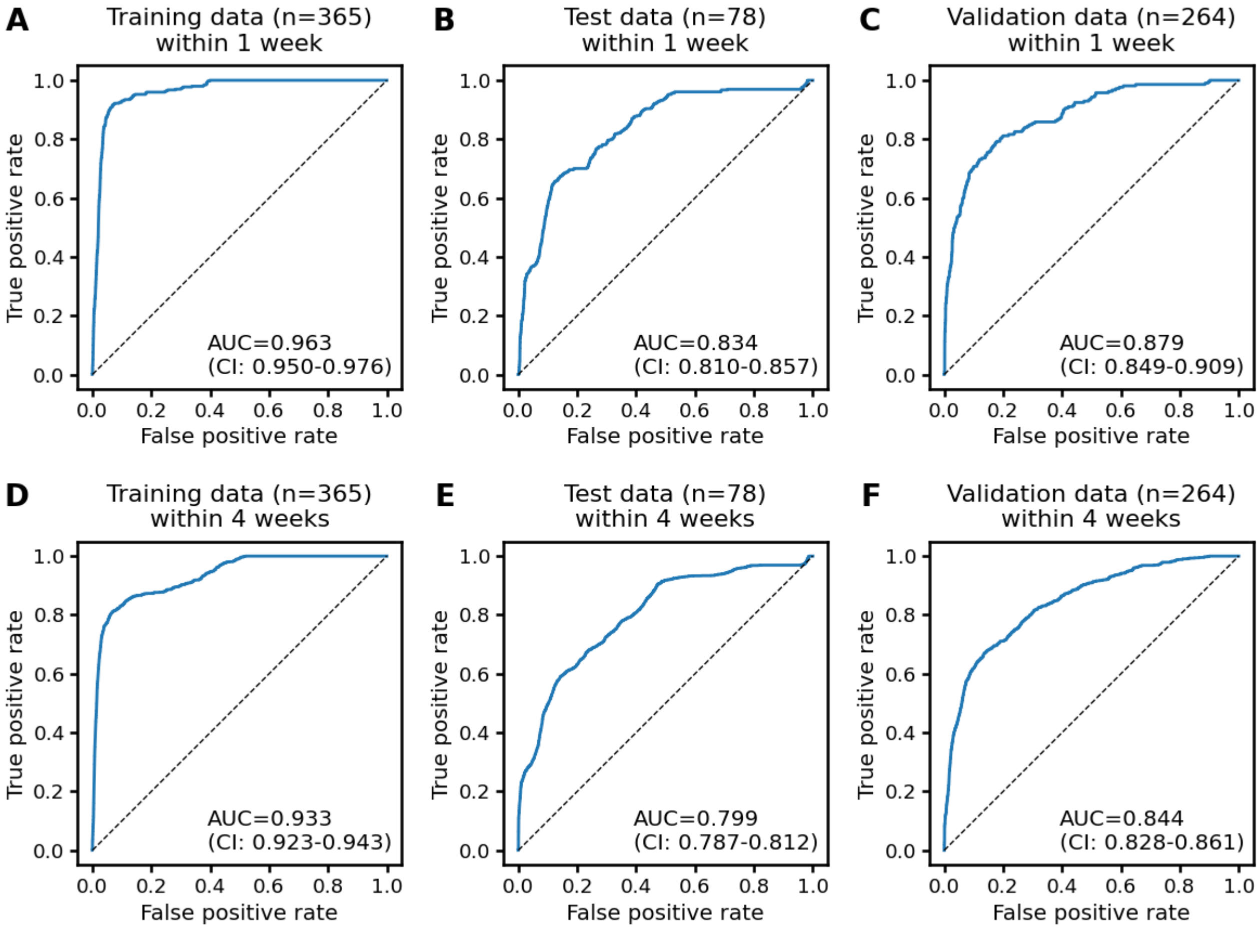


Supplementary Figure 3. Receiver operating characteristic curves for predictions within one week or four weeks. (A) Training data, within one week. (B) Test data, within one week. (C) Validation data, within one week. (D) Training data, within four weeks. (E) Test data, within four weeks. (F) Validation data, within four weeks. AUC: area under the curve; CI: confidence interval.

Supplementary Table 1. Antihypertensive medication use.

| Patient no | Cohort | Classification based on HBPM | HDP onset[1] (days of gestation) | Delivery (days of gestation) | Start of treatment (days of gestation) | Medication |
|---|---|---|---|---|---|---|
| 1 | Development | HDP | 256 | 304 | 297 | Nifedipine |
| 2 | Development | HDP | 260 | 296 | 280 | Nifedipine |
| 3 | Development | HDP | 244 | 286 | 277 | Nifedipine |
| 4 | Development | HDP | 251 | 277 | 267 | Nifedipine |
| 5 | Development | HDP | 246 | 273 | 257 | Nifedipine |
| 5 | Development | HDP | 246 | 273 | 257 | Methyldopa |
| 6 | Development | HDP | 249 | 287 | 262 | Labetalol |
| 7 | Validation | NBP | n/a | 270 | 275 | Amlodipine |
| 8 | Validation | HDP | 249 | 269 | 262 | Nifedipine |
| 8 | Validation | HDP | 249 | 269 | 273 | Amlodipine |
| 9 | Validation | HDP | 247 | 262 | 261 | Nifedipine |
| 10 | Validation | NBP | n/a | 269 | 275 | Amlodipine |

HBPM: home blood pressure measurement; HDP: hypertensive disorders of pregnancy; NBP: normal blood pressure.

[1]HDP onset was defined based on HBPM.

Supplementary Table 2. List of candidate features.

| Feature name |
|---|
| Mean SBP |
| Mean DBP |
| Mean HR |
| Standard deviation of SBP |
| Standard deviation of DBP |
| Standard deviation of HR |
| Slope of SBP against day |
| Slope of DBP against day |
| Slope of HR against day |
| Correlation coefficient between SBP and DBP |
| Correlation coefficient between SBP and HR |
| Correlation coefficient between DBP and HR |

SBP: systolic blood pressure; DBP: diastolic blood pressure; HR: heart rate.

Supplementary Table 3. Prediction performance of the proposed method in a sensitivity analysis.

| | Sensitivity | Specificity | PPV | NPV | PLR | NLR |
|---|---|---|---|---|---|---|
| Development cohort, training (n=365; NBPs: 339; HDPs: 26) | 0.633 0.499-0.754) | 0.921 (0.885-0.949) | 0.613 (0.481-0.734) | 0.927 (0.892-0.954) | 8.049 (5.238-12.37) | 0.398 (0.285-0.556) |
| Development cohort, test (n=78; NBPs: 39; HDPs: 39) | 0.667 (0.505-0.804) | 0.944 (0.813-0.993) | 0.933 (0.779-0.992) | 0.708 (0.559-0.830) | 12.00 (3.068-46.93) | 0.353 (0.228-0.545) |
| Validation cohort, Toyama (n=119; NBPs: 98; HDPs: 21) | 0.612 (0.462-0.748) | 0.886 (0.787-0.949) | 0.789 (0.627-0.904) | 0.765 (0.658-0.852) | 5.357 (2.689-10.67) | 0.438 (0.305-0.629) |
| Validation cohort, Miyagi (n=145; NBPs: 133; HDPs: 12) | 0.507 (0.382-0.632) | 0.910 (0.824-0.963) | 0.829 (0.679-0.928) | 0.683 (0.584-0.771) | 5.655 (2.684-11.91) | 0.541 (0.420-0.697) |
| Validation cohort, all (n=264; NBPs: 231; HDPs: 33) | 0.552 (0.457-0.644) | 0.899 (0.838-0.942) | 0.810 (0.706-0.890) | 0.719 (0.648-0.782) | 5.444 (3.279-9.038) | 0.499 (0.405-0.615) |

In this sensitivity analysis, hypertension was redefined using HBP criteria for the general population: (1) HBP ≥ 135/85 mmHg was observed twice in a row within 48 hours, or (2) HBP ≥ 155/105 mmHg was observed once. The 95% confidence intervals are in parentheses. HBP: home blood pressure; PPV: positive predictive value; NPV: negative predictive value; PLR: positive likelihood ratio; NLR: negative likelihood ratio.

Supplementary Table 4. Prediction performance of the baseline method.

| | Sensitivity | Specificity | PPV | NPV | PLR | NLR |
|---|---|---|---|---|---|---|
| Development cohort, training (n=365; NBPs: 339; HDPs: 26) | 0.385 (0.202-0.594) | 0.903 (0.866-0.932) | 0.233 (0.118-0.386) | 0.950 (0.921-0.971) | 3.951 (2.203-7.088) | 0.682 (0.502-0.926) |
| Development cohort, test (n=78; NBPs: 39; HDPs: 39) | 0.282 (0.150-0.449) | 0.923 (0.791-0.984) | 0.786 (0.492-0.953) | 0.562 (0.433-0.686) | 3.667 (1.108-12.14) | 0.778 (0.626-0.966) |
| Validation cohort, Toyama (n=119; NBPs: 98; HDPs: 21) | 0.143 (0.030-0.363) | 0.888 (0.808-0.943) | 0.214 (0.047-0.508) | 0.829 (0.743-0.895) | 1.273 (0.389-4.169) | 0.966 (0.800-1.166) |
| Validation cohort, Miyagi (n=145; NBPs: 133; HDPs: 12) | 0.250 (0.055-0.572) | 0.932 (0.875-0.969) | 0.250 (0.055-0.572) | 0.932 (0.875-0.969) | 3.694 (1.152-11.85) | 0.804 (0.578-1.119) |
| Validation cohort, all (n=264; NBPs: 231; HDPs: 33) | 0.182 (0.070-0.355) | 0.913 (0.869-0.946) | 0.231 (0.090-0.436) | 0.887 (0.839-0.924) | 2.100 (0.910-4.846) | 0.896 (0.759-1.057) |

In this sensitivity analysis, hypertension was redefined using HBP criteria for the general population: (1) HBP ≥ 135/85 mmHg was observed twice in a row within 48 hours, or (2) HBP ≥ 155/105 mmHg was observed once. The 95% confidence intervals are in parentheses. HBP: home blood pressure; PPV: positive predictive value; NPV: negative predictive value; PLR: positive likelihood ratio; NLR: negative likelihood ratio.
